# Evaluating the impact of curfews and other measures on SARS-CoV-2 transmission in French Guiana

**DOI:** 10.1101/2020.10.07.20208314

**Authors:** Alessio Andronico, Cécile Tran Kiem, Juliette Paireau, Tiphanie Succo, Paolo Bosetti, Noémie Lefrancq, Mathieu Nacher, Félix Djossou, Alice Sanna, Claude Flamand, Henrik Salje, Cyril Rousseau, Simon Cauchemez

## Abstract

While general lockdowns have proven effective to control SARS-CoV-2 epidemics, they come with enormous costs for society. It is therefore essential to identify control strategies with lower social and economic impact. Here, we report and evaluate the control strategy implemented during a large SARS-CoV-2 epidemic in June-July 2020 in French Guiana that relied on curfews, targeted lockdowns and other measures. We find that the combination of these interventions reduced the basic reproduction number of SARS-CoV-2 from 1.7 to 1.1, which was sufficient to avoid saturation of hospitals. We estimate that thanks to the young demographics across the territory, the risk of hospitalisation following infection was 0.3 times that of metropolitan France and that about 20% of the population was infected by July. Our model projections are consistent with a recent seroprevalence study. The study showcases how mathematical modeling can be used to support healthcare planning and decision making in a context of high uncertainty.

## Introduction

Following its emergence in China in December 2019, the SARS-CoV-2 virus has quickly spread around the world [1]. To avoid saturation of their healthcare systems, many countries enforced nationwide lockdowns. While such an approach has demonstrated its efficacy for transmission control [2–5], it comes with very high social and economic costs [6,7]. As a consequence, lockdowns cannot be sustained for long periods of time and it remains essential to identify sets of interventions with lower impact on society that are effective enough to contain epidemic rebounds of SARS-CoV-2.

While mathematical modelling can help evaluate the likely impact of different strategies, demonstration of efficacy comes when these approaches are successfully implemented in the field. However, testing new control strategies during an ongoing SARS-CoV-2 outbreak is a difficult and stressful exercise for policy makers. If the new strategy does not reduce transmission rates sufficiently, this could have dramatic consequences for the healthcare system. Furthermore, underlying delays between infection and hospital admission mean that the effect of the interventions on the healthcare system can only be confidently measured a few weeks after their implementation. During this time period, policy makers do not know whether the strategy will be successful, case counts may continue to rise along with the pressure on the healthcare system and the time window to implement corrective measures is shrinking. Confronted with such an uncertain and quickly evolving situation, it is essential to plan for the worst. In this context, mathematical modelling can help construct realistic scenarios to inform plans for a surge in ICU capacity or a strengthening of control measures [4,8–10] and quickly revise these scenarios as more data become available.

Ultimately, the insight we gain from these local experiences will be key to progressively optimize control strategies and determine the minimal set of interventions that is sufficient for the control of SARS-CoV-2 epidemic rebounds. Here, we report on the local experience of French Guiana, a French overseas territory located in Latin America in the Amazonian forest complex, where authorities managed to contain a large SARS-CoV-2 epidemic with the use of curfews, local lockdowns and other measures. We describe the epidemic dynamics, the interventions that were implemented and their impact and show how mathematical modelling was used throughout the outbreak to support healthcare planning and policy making in a context of high uncertainty.

## Results

### Epidemic of SARS-CoV-2 in a young population: impact on healthcare demand

The severity of SARS-CoV-2 infection increases with the age of the individual [2,11,12]. As a consequence, the impact of a SARS-CoV-2 epidemic on the healthcare system is expected to vary with the demographic structure of the population it is spreading in [13]. The population of French Guiana is substantially younger than that in metropolitan France, with a median age of 27y in French Guiana compared to 42y in metropolitan France (Figure 1B). We used our mathematical model [2] to anticipate how these differences were expected to affect stress on the healthcare system for a given level of circulation of the virus.

**Figure 1:**
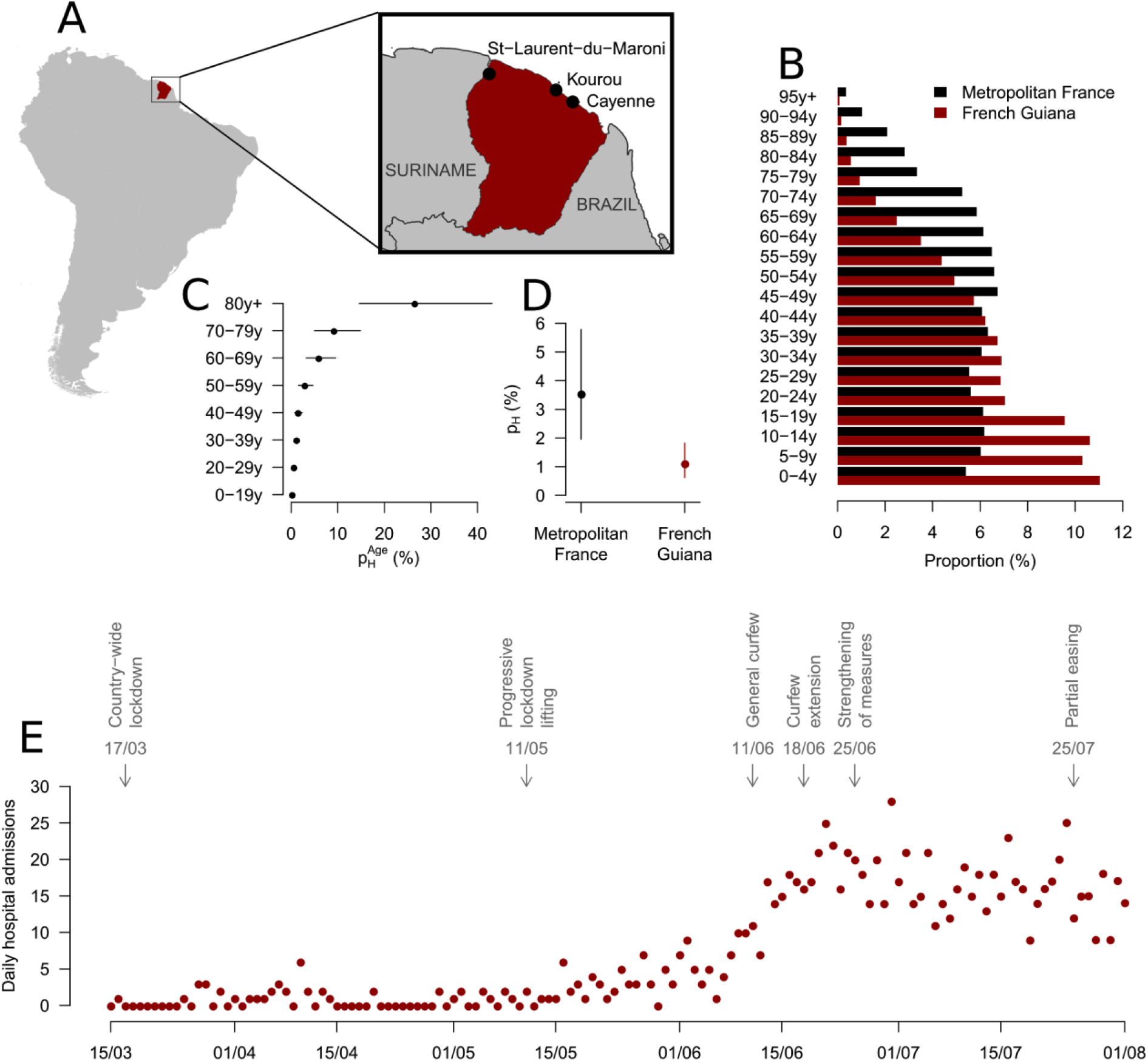
**A**. Map of French Guiana. **B**. Population pyramids for Metropolitan France and French Guiana. **C**. Age-specific probability of hospitalization given infection *p*_*H*_ ^*Age*^ in Metropolitan France (%). **D**. Average probability of hospitalization given infection *p*_*H*_ in Metropolitan France and French Guiana (%). **E**. Daily hospital admissions in French Guiana and timeline of interventions.

During the first pandemic wave in metropolitan France, we estimated age-specific probabilities of hospitalisation given infection under the assumption that children were half as infectious as adults (Figure 1C) [2]. We found that, on average, in metropolitan France, a person infected by SARS-CoV-2 had 3.5% [1.9%, 5.8%] probability of being hospitalized (Figure 1D) [2]. Applying these age-specific probabilities to the demographic structure and expected contact patterns in French Guiana, we anticipated that the average probability of hospitalization upon infection would be 1.1% [0.6%, 1.8%] in this population (Figure 1D). This means that, for the same number of infections, we expected 0.32 times as many hospitalisations in French Guiana as in metropolitan France.

### Measures implemented to control the spread of SARS-CoV-2 in French Guiana

Like the rest of France, a territory-wide lockdown was imposed in French Guiana from March 17th 2020 to May 11th 2020. At that point, the lockdown was eased but not entirely lifted: schools, places of worship and movie theaters stayed closed, while restaurants and bars were allowed to reopen but were limited to outdoor sitting for on-site dining. A curfew was established from 11PM to 5AM every day except in Saint Georges, a city located on the border with Brazil, where a complete lockdown was maintained. Terrestrial borders were closed and travel restrictions were implemented to reduce the risk of spatial spread. Despite these measures, and concomitantly with the acceleration of the epidemic in Brazil (that neighbors French Guiana), the number of cases started to rise at the end of May 2020. In response to this rise, control measures were strengthened on June 10th, with the general curfew being extended from 9PM to 5AM during weekdays and for the entire day on Sundays. On June 18th, the curfew was extended again from 7PM to 5AM during weekdays and from 3PM on Saturday during the weekend. Additional measures from June 25th, included the start of the curfew from 5PM during weekdays and from 1PM on Saturday during the weekend, enforced closure of all restaurants, the closure of the Brazilian border and the lockdown of 23 high-risk areas. There were also important screening campaigns with more than 1300 tests per 100 000 inhabitants per week in the weeks following June 15^th^ 2020. A partial easing of the more stringent measures - in particular the local lockdowns - took place on July 25th (Figure 1E). The implementation and schedule of the curfews were adapted to each area according to the epidemiological situation.

### Planning for the worst

A key challenge for the management of such an epidemic is that it is not possible to evaluate the impact of new control measures on hospital admissions for the 2-3 weeks that follow their implementation (about 11 days from infection to hospitalisation [14] and between 5 and 10 days to accumulate sufficient data to characterize trends post intervention). During this stressful period, cases continue to rise and we do not know if control measures will provide the expected reduction in infections. At a time of such high uncertainty, it is important to plan adequate healthcare capacity and a potential strengthening of control measures in case transmission rates following intervention are not sufficiently reduced. We ran two analyses during this period to support such planning.

#### What if control measures do not reduce transmission?

To support healthcare planning and ensure appropriate scaling of the local ICU capacity, we evaluated healthcare demand in a worst-case scenario assuming that local transmission rates would remain unchanged despite the additional control measures. This was done considering a broad spectrum of scenarios for the probability of hospitalisation of an infected individual (baseline: *p*_*H*_= 1.1%; low: *p*_*H*_= 0.6%; high: *p*_*H*_= 1.8%) (Figure 1D) and the duration of stay in ICU (baseline: *τ*_*ICU*_ = 11.4 days; short: *τ*_*ICU*_ = 8.0 days; long: *τ*_*ICU*_ = 15.0 days) based on early estimates from local data.

Calibrated to data available on June 18th, our model identified that there had been an acceleration of the epidemic around May 20th, with the basic reproduction number increasing from 1.35 [1.26, 1.45] (before May 20th) to 1.78 [1.68, 1.88] (afterwards). In the scenario where the transmission rate remained unchanged following interventions, the peak of the epidemic was expected in July (Figure 2A and 2B). Depending on the severity scenario, the peak number of daily hospitalisations was projected at 48 [34, 65] for average severity (low severity: 28 [18, 40]; high severity: 75 [55, 96]) while the peak number of daily ICU admissions was projected at 11 [4, 18] (low severity: 6 [2, 11]; high severity: 16 [8, 25]). The number of general ward beds required at the peak was estimated at 454 [383, 528] for average severity (low severity: 262 [221, 305]; high severity 715 [592, 831]), and the number of ICU beds at 110 [86, 137] (low severity: 63 [47, 82], high severity: 173 [135, 210]). Figure S1 shows that the projected timing and intensity of the peak were overall more sensitive to the probability of hospitalisation *p*_*H*_ than to the duration of stay in ICU *τ*_*ICU*_.

**Figure 2:**
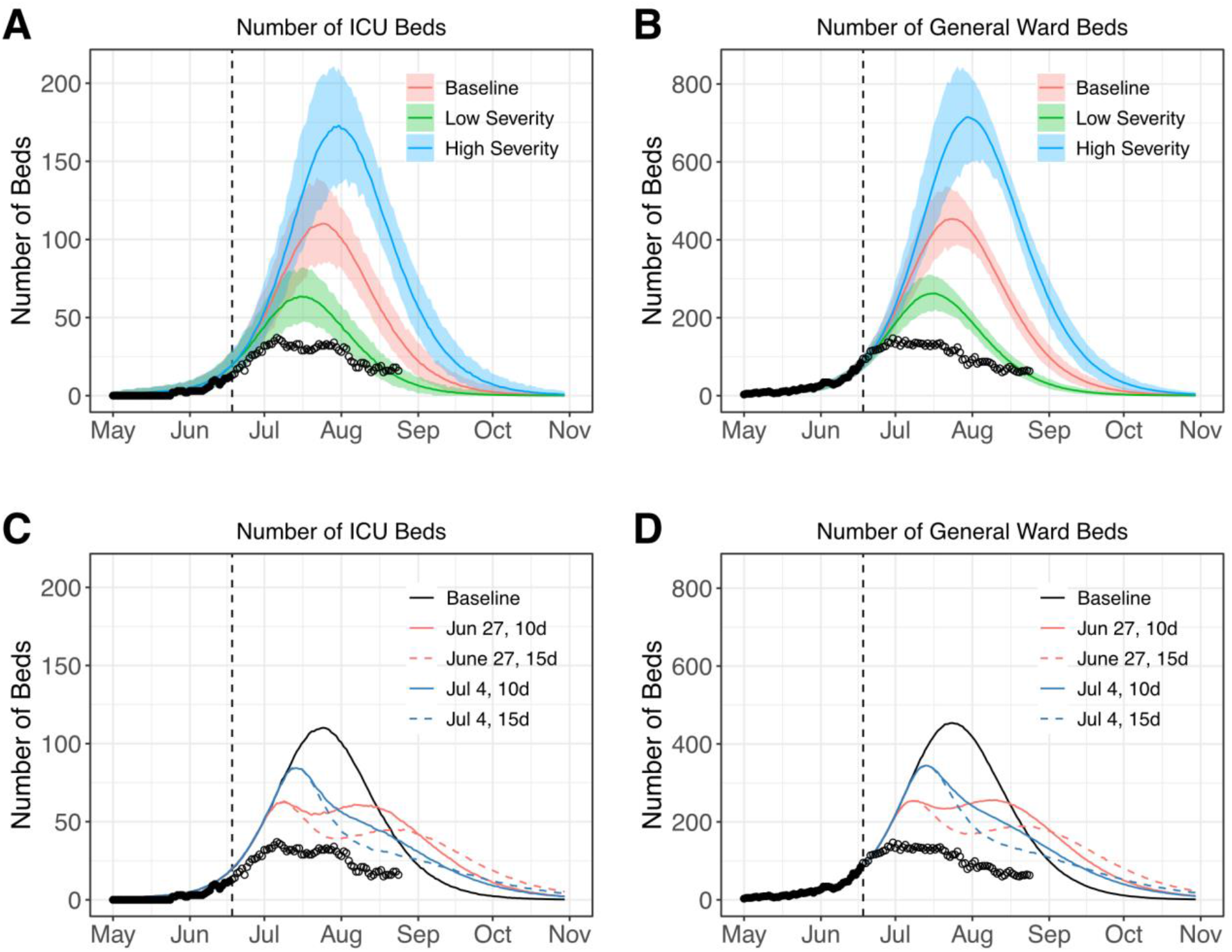
Analyses made on June 18th 2020 describing a worst-case scenario with no change in transmission rates and the impact of a short-term lockdown. **A. and B**. Projections for the number of ICU and general ward beds required under different severity scenarios (baseline in red, low severity in green, high severity in blue). Solid lines indicate model posterior means while color areas indicate 95% credible intervals. **C. and D**. Projections for the average number of ICU and general ward beds required under different territory-wide lockdown scenarios (black represents our baseline model, red represents lockdowns starting on June 27th, blue represents lockdowns starting on July 4th, solid lines correspond to lockdowns lasting for 10 days, while dashed lines correspond to lockdowns lasting for 15 days). In all panels, black dots indicate data used to calibrate the models, while empty circles denote data not available at the time of the analyses. The dashed line in all panels indicates the date of the analyses (June 18th).

#### Impact of a short lockdown on peak intensity

Given the quickly expanding epidemic, the limited ICU capacity and the possibility that existing control measures might not sufficiently reduce transmission, the option of imposing a short lockdown in French Guiana was also considered by French authorities. We assessed how such a lockdown might ease peak healthcare demand, assuming that it would lead to similar transmission rates as those observed in metropolitan France with a lockdown reproduction number of 0.7. This was done considering different start dates and durations for the lockdown (Figure 2C and 2D). We found that a territory-wide 10 days lockdown would reduce the required number of general ward beds from 454 [383, 528] to 256 [222, 290] (if started on June 27th) or to 345 [276, 427] (if started on July 4th). Similarly, the expected number of required ICU beds was projected to decrease from 110 [86, 137] to 63 [45, 86] (lockdown starting on June 27th) or to 84 [61, 111] (lockdown starting on July 4th).

Eventually given the societal and economic cost associated with a territory-wide lockdown, this strategy was ruled out and it was decided to implement less drastic measures accompanied by an increase of ICU bed capacity and the planning of patient transfers to hospitals in Martinique and Guadeloupe.

### Initial estimates of the impact of interventions on the epidemic trajectory

Once sufficient time had passed, we ascertained the impact of the new interventions by adding a change point, to be estimated, for the transmission rate (model M2). From June 27th onwards, this model had better DIC support than model M1 with no change in transmission (Figure S2). Model M2 estimated that the transmission rate was reduced from June 15th [10th, 19th], coincidentally with the strengthening of control measures (Figure S3). According to the model, the basic reproduction number went from 1.40 [1.32, 1.49] before May 20th to 1.71 [1.65, 1.77] between May 20th and June 15th and 1.14 [0.95, 1.31] after June 15th (Figure 3A). This suggests that the strict curfew measures were successful at reducing transmission. With these reduced transmission rates, projections of the number of hospital and ICU beds required at the peak dropped to 28 [17, 42] ICU beds and 162 [127, 203] general ward beds (Figure 3C and 3D).

**Figure 3:**
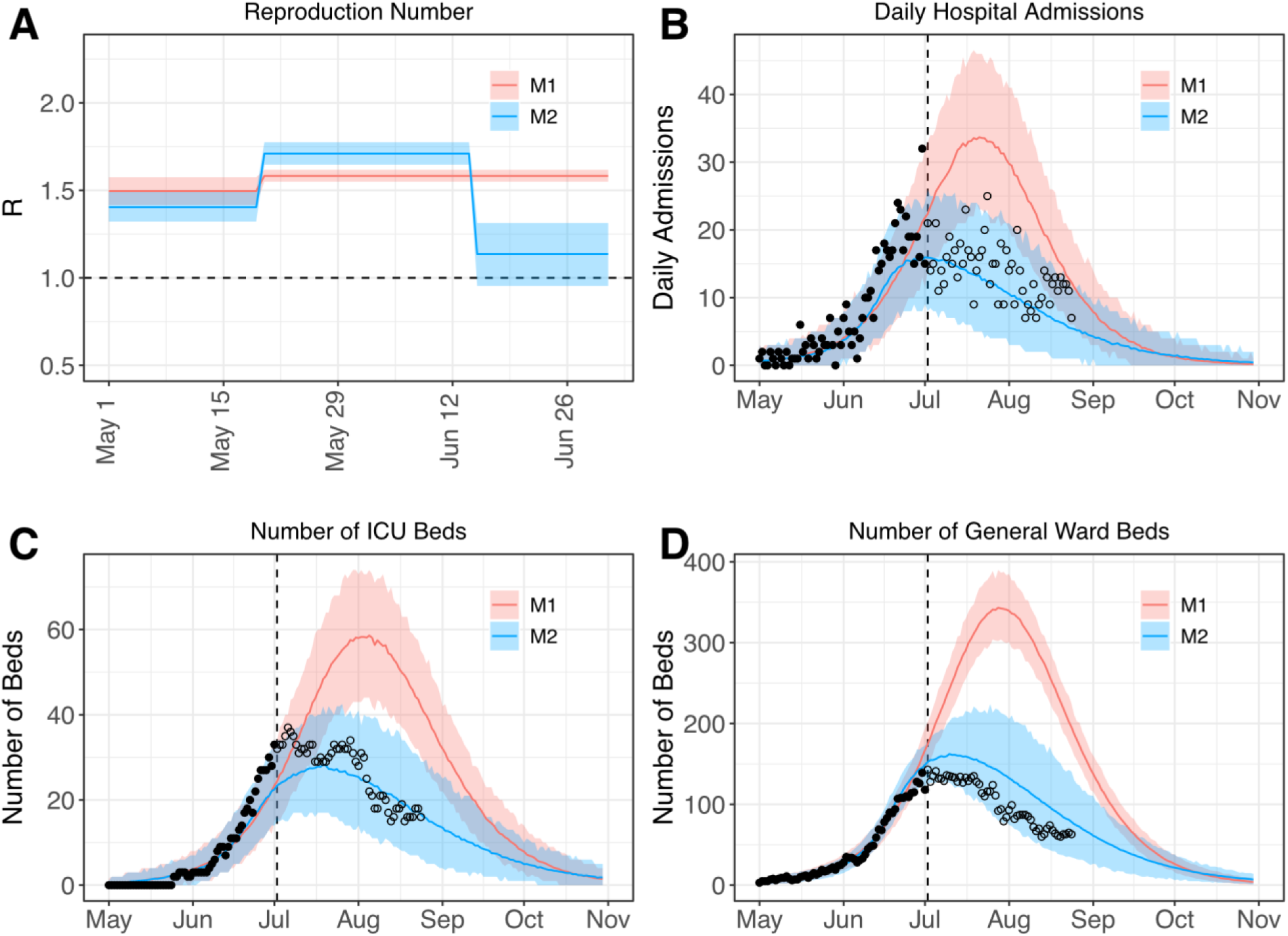
Analyses made on July 2nd 2020 evaluating the impact of control measures implemented in French Guiana on transmission and healthcare demand. **A**. Estimated reproduction number through time. **B. to D**. Projections for the number of daily hospital admissions (**B**) and ICU (**C**) and general ward (**D**) beds. Solid lines indicate model posterior means while color areas indicate 95% credible intervals. Red is used for model M1 (one change point for the transmission rate), while blue is used for model M2 (two change points for the transmission rate). In all panels black dots indicate data used to calibrate the models, while empty circles denote data not available at the time of the analyses. The dashed line panels B-D indicates the date of the analyses (July 2nd).

#### Latest assessments, model validation and improvements

Figure 4 shows the projections obtained from model M2 calibrated using data available on August 25th 2020. The probability *p*_*ICU*_ of ICU admission given hospitalisation was estimated at 15.7% [13.9%, 17.6%] and the time spent in ICU *τ*_*ICU*_ at 15.0 [13.1, 17.4] days. Our projections have been relatively stable since the beginning of July (Figure 4A-D).

**Figure 4:**
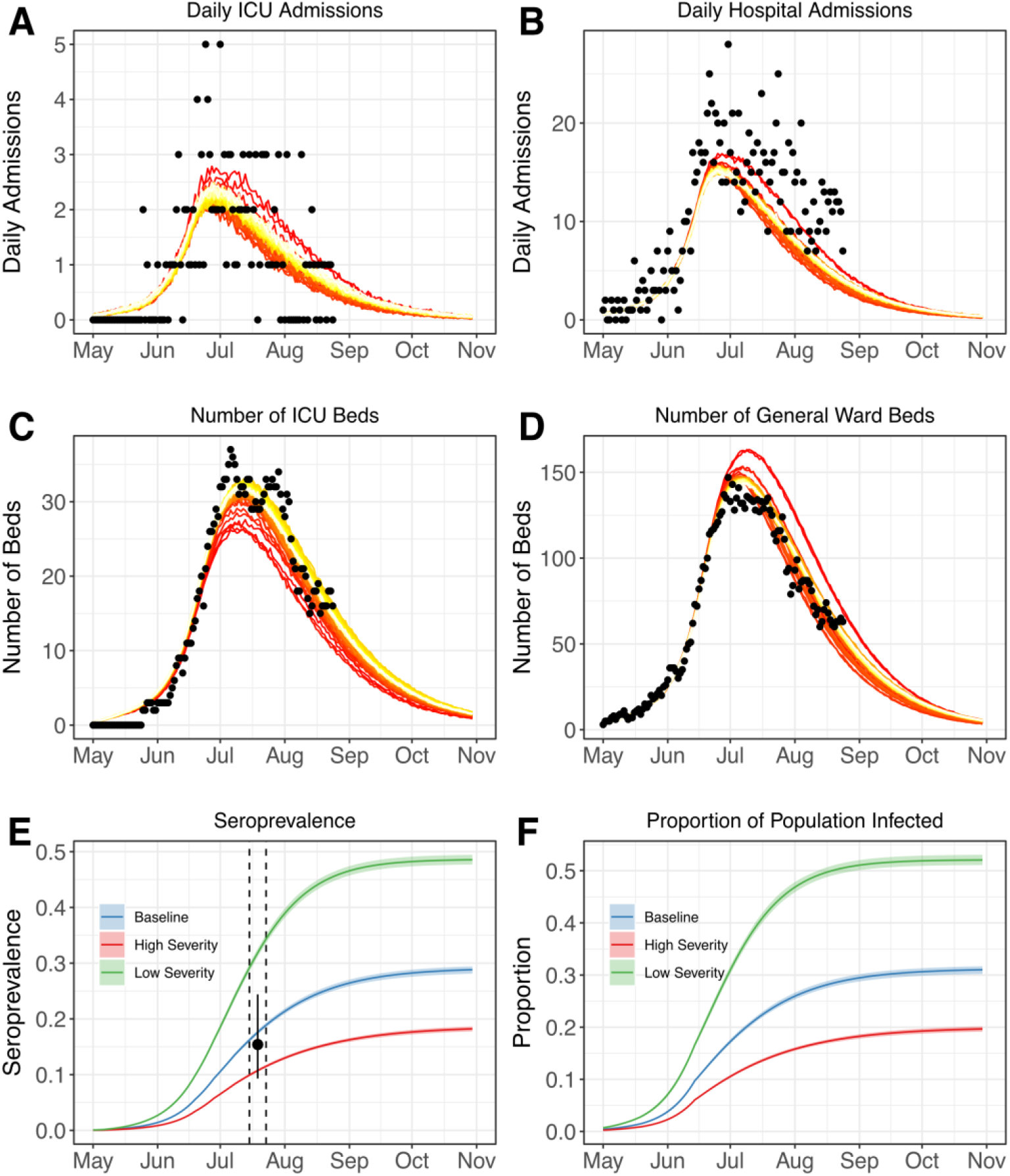
Analyses made on August 25th 2020. **A. to D**. Projections for the number of daily ICU (**A**) and hospital admissions (**B**), ICU (**C**) and general ward (**D**) beds. **(E)**. Projections for the seroprevalence measured with the Euroimmun assay. Solid lines indicate model posterior means while color areas indicate 95% credible intervals. The black dot indicates the seroprevalence estimated in [15] between 15-23 July 2020 (dashed lines). **(F)**. Projections for the proportion infected. Solid lines indicate model posterior means while color areas indicate 95% credible intervals. In panels A to D, darker red colors correspond to older projections while lighter yellow colors correspond to more recent ones (from July 1st to August 26th). Black dots indicate actual data.

We retrospectively validated our model by comparing model predictions with the results of a seroprevalence survey performed between July 15th and July 23rd with the Euroimmun assay [15]. Assuming a time-dependent assay sensitivity (0% during incubation, 30.3% up to 10 days after symptom onset, 75% between 10 and 20 days after symptom onset, and 93.8% afterwards) as per the distributor specification, our model estimated that 17.6% [17.2%, 18.0%] of the population was seropositive for SARS-CoV-2 between July 15th and July 23rd for average severity (low severity: 31.8% [31.1%, 32.6%]; high severity: 10.7% [10.5%, 11.0%]) (Figure 4E). Estimates for average severity are close to the seroprevalence of 15.4% [9.3%, 24.4%] obtained in the serosurvey, indicating that our average severity scenario remains the one that is best supported by the data. Projecting forward, we anticipate that 30.6% [29.9%, 31.3%] (low severity: 51.9% [50.9%, 52.9%]; high severity: 19.3% [18.8%, 19.7%]) of the population in French Guiana will have been infected by October 1st 2020 (Figure 4F).

## Discussion

In this paper, we characterized the epidemic dynamics of SARS-CoV-2 in French Guiana, evaluated the impact of control measures that were implemented to contain a large SARS-CoV-2 epidemic there, and described how mathematical modelling was used during this crisis to support policy making and planning.

The nation-wide lockdown that was implemented across France from March 17th 2020 to May 11th 2020 likely prevented a surge of SARS-CoV-2 infections in French Guiana during this period. However, while a number of control measures remained in place in French Guiana after the lockdown, they were insufficient to stop an important epidemic rebound. This epidemic was likely facilitated by the proximity of Brazil, a country that has experienced a very important pandemic wave [16,17], notably in neighboring Amazonian states. Confronted with an important surge in COVID-19 cases, French authorities implemented a set of strong measures including curfews and localized lockdowns. During curfews, individuals can go to work and live a relatively normal life during the day, but social interactions are limited in the evenings and weekends. This approach therefore targets social interactions in the private sphere where social distancing is more likely to be relaxed. While smaller than that of a full lockdown, the economic impact of a curfew remains important in particular for the hospitality, catering and recreational sectors, as well as for a large part of the undeclared jobs on which the most precarious rely on in French Guiana.

We estimate that, added to existing measures, these interventions further reduced the basic reproduction number by 36% from 1.7 (prior to interventions) to 1.1 (following implementation). This change in epidemic dynamics strongly reduced predicted ICU beds needs for the epidemic peak from 110 to 32, thereby avoiding saturation of ICUs. The territory was also able to manage the influx of patients thanks to an expansion of ICU capacity (from 11 on May 1 2020 to 54 on July 22 2020) and the transfer of 7 ICU patients to Martinique and 6 to Guadeloupe, two French overseas territories located in the Caribbean.

In agreement with a seroprevalence study [15], we find that the infection attack rate of SARS-CoV-2 in French Guiana is currently one of the highest in France, likely higher than that estimated for Grand Est (7.7%-10.2% between May 4th and June 22nd) and Île-de-France (Paris area) (9.1%-10.9% between May 4th and June 22nd), the two regions of metropolitan France that have been the most affected by the first pandemic wave [18]. This may seem surprising since the impact of the epidemic on hospitalisations and deaths was substantially lower in French Guiana (183 hospitalisations by July 1st and 13 deaths per 100,000 inhabitants by July 18th) than in Grand Est (276 hospitalisations by May 25th and 60 deaths per 100,000 inhabitants by June 11th) and Île-de-France (280 hospitalisations by May 25th and 56 deaths per 100,000 inhabitants by June 11th). This apparent discrepancy was anticipated by our model and can be explained because the population of French Guiana is substantially younger than that of metropolitan France (Figure 1). This shows that it is essential to account for the age structure of a population to properly evaluate the impact of SARS-CoV-2 on its healthcare system. In an older population, it is likely that pressure on the healthcare system would have occurred earlier in the epidemic, leading to earlier implementation of control measures and lower seroprevalence. Improvements in patient management thanks for example to anticoagulation, steroid and ventilation may have also contributed to averting deaths [19,20].

Major methodological developments have been made in the last few years to strengthen epidemic forecasting, with seasonal influenza or dengue constituting good case studies [21,22]. In a typical seasonal influenza epidemic, measures to reduce transmission in the general population are limited. As a consequence, once the epidemic has started, we expect that it will follow its natural course and that its trajectory can be forecasted if we have a good understanding of its key drivers (e.g. impact of the climate, population immunity, school holidays and circulating influenza subtype). In contrast, for SARS-CoV-2, unprecedented control measures are being implemented to limit spread; in addition, individuals are likely to naturally modify their behaviours (e.g. to reduce their contacts) as the pandemic progresses in their community [23]. A simple international comparison shows how the control measures and behaviours that are adopted can radically change the course of the pandemic from scenarios of near-suppression in South Korea and New Zealand to much less favorable ones in Brazil and the US. In addition, both control measures and individual behaviours may quickly change with the epidemiological situation, in a way that may be hard to anticipate. All these elements explain why it is much more challenging to forecast the trajectory of the SARS-CoV-2 pandemic wave than that of, for example, a seasonal influenza epidemic. Given these difficulties, we prefer to talk about scenario analysis rather than forecasts.

French Guiana constitutes an interesting case study where a combination of strict interventions including curfews and localized lockdown substantially reduced SARS-CoV-2 transmission. We need to build on these local experiences to progressively determine the optimal set of interventions required to contain SARS-CoV-2 pandemic waves.

## Methods

We used a deterministic mathematical model to describe the transmission of SARS-CoV-2 and subsequent disease progression in the population of French Guiana. The compartmental structure of the model closely followed our previous work [2]: upon infection, susceptible individuals enter a first latent compartment where they are not infectious, while a second exposed compartment is used to capture individuals who are infectious but not yet symptomatic. Once infected, individuals can develop severe disease and require hospital and/or ICU care. We used two versions of the model. A first version explicitly accounted for the age structure in the population. To describe contact patterns in the population of French Guiana, we used a contact matrix from Suriname [24], a neighboring territory with similar population structure. We adjusted the contact matrix for age-groups 0-9y, 10-19y, 20-29y, 30-39y, 40-49y, 50-59y, 60-69y, 70y+ accounting for the population structure of French Guiana. In order to accelerate computation and shorten the turnaround time of our analyses, we developed a second version of the model in which we no longer explicitly included the population age structure in the model. We instead relied on a single severity parameter, the average probability of hospitalization given infection *p*_*H*_. Assuming that the probability of infection is proportional to the daily number of contacts within each age group (*C*_*i*_ for age-group i) [2], this severity parameter can be estimated from the age-specific probability of hospitalization upon infection (*p*_*H*_^*i*^ for age-group i) and the age distribution of the target population as follows:

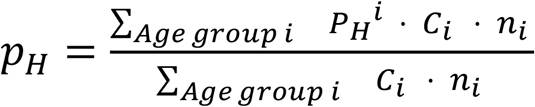 where *n*_*i*_ is the number of individuals aged i in the target population. Throughout this analysis, we considered young people to be half as infectious as adults. Results obtained with the full age-structured model under our final assumptions closely matched those obtained with the simpler version (Figure S4).

In order to capture trends in the epidemic trajectory following the strengthening of control measures in French Guiana, we modified the structure of our model for the analyses we performed at the beginning of July 2020: while our initial model (M1) had a single change point for the transmission rate, our final model (M2) had two change points for this parameter. Table S1 summarizes the models’ key parameters.

The simulations were seeded on April 21st 2020 with an initial number of infectious individuals - split into the exposed and infectious compartments proportionally to the time spent in each compartment - that was estimated jointly with the parameters in Table S1.

We fitted our models to daily hospitalization count data extracted from the SI-VIC database, which stores data on COVID-19 patients hospitalized in public and private hospitals in metropolitan France and overseas French territories. The data were corrected for reporting delays as previously described [2].

The model parameters were estimated via Markov Chain Monte Carlo (MCMC) sampling assuming a Poisson observation process and using uniform, non-informative, priors. We relied on the Deviance Information Criterion (DIC) for model comparison and selection [25], with smaller DIC values indicating stronger support for the model.

## Data Availability

All data will be made available.

