## Supplementary figures and table for "Evaluating the impact of curfews and other measures on SARS-CoV-2 transmission in French Guiana"

### Supplements

**Figure S1: Sensitivity to severity and duration of stay in ICU. A.** Projected number of ICU beds according to different severity scenarios: baseline ($p_H$= 1.1%, red), low ($p_H$= 0.6%, green), and high ($p_H$= 1.8%, blue). **B.** Projected number of ICU beds according to different durations of stay in ICU: baseline ($\tau_{ICU}$= 11.4 days, red), short ($\tau_{ICU}$= 8.0 days, brown), and long ($\tau_{ICU}$= 15.0 days, purple). Black dots indicate data used to calibrate the models, while empty circles denote data not available at the time of the analyses.

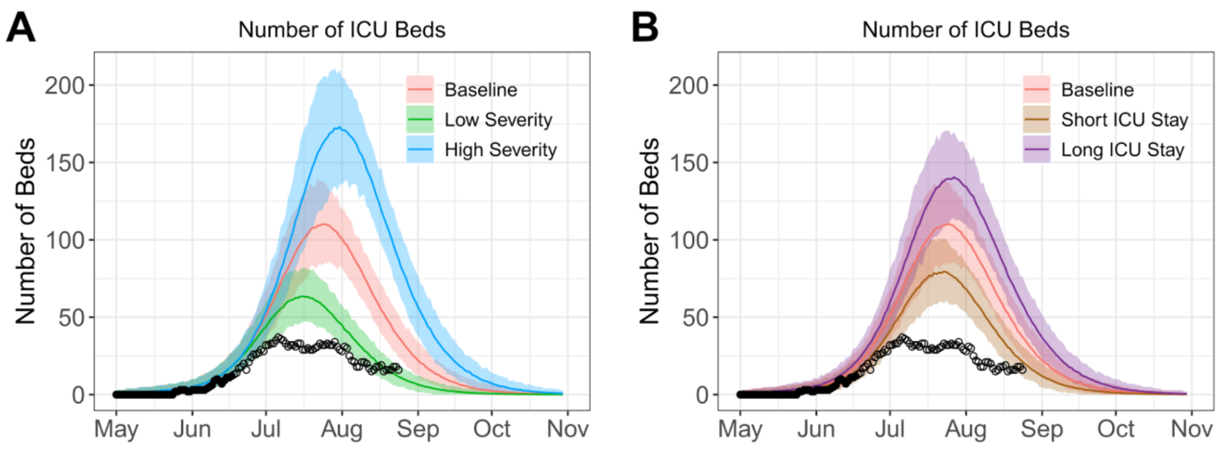

**Figure S2: Model selection.** DIC difference (DIC of model M1 - DIC of model M2) for models calibrated from June 19th to June 29th. The dashed lines indicate a DIC difference of 4 units. Model M2 has a change point on June 15th (Figure S3).

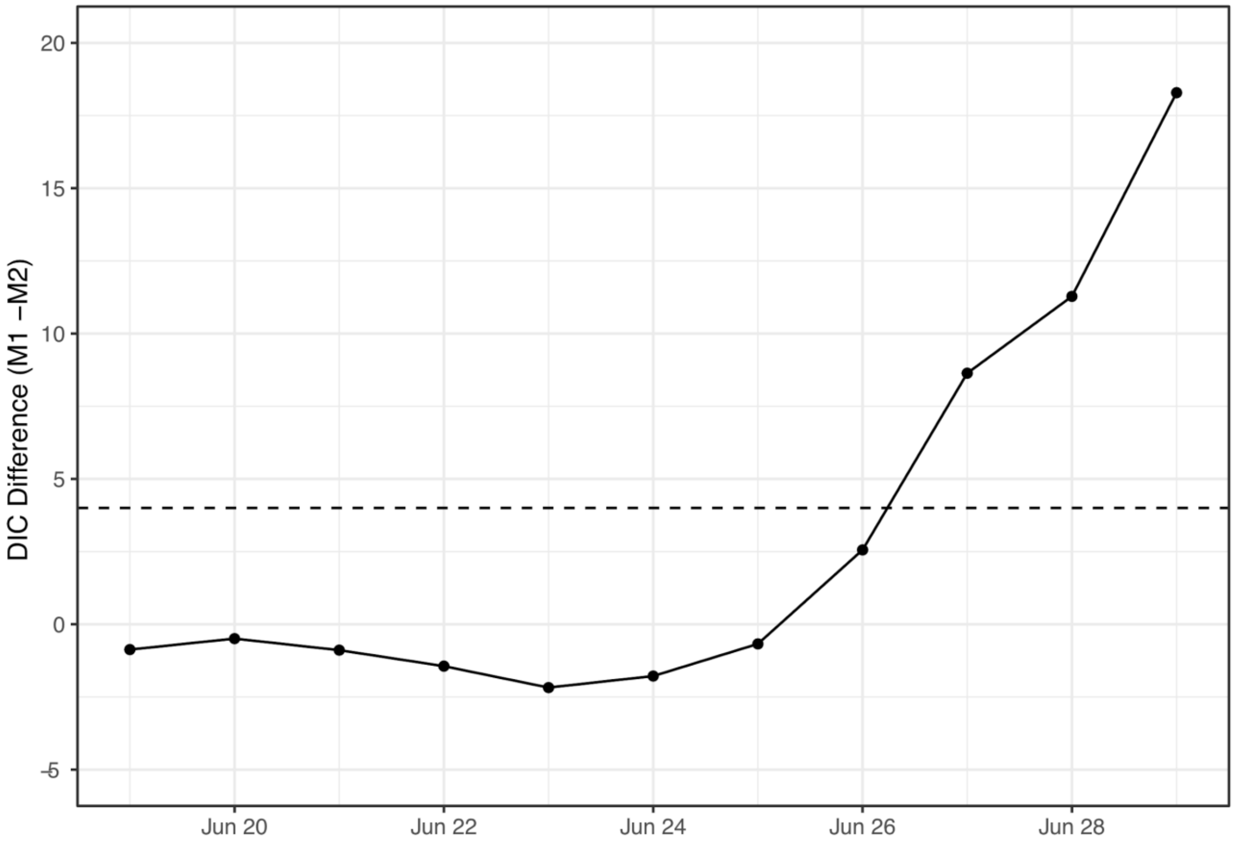

**Figure S3: Choice of transmission rate change point for model M2.** Model DIC for change points ranging from June 6th to June 26th. The lowest DIC is obtained for a change point on June 15th. The dashed lines indicate a DIC difference of 4 units.

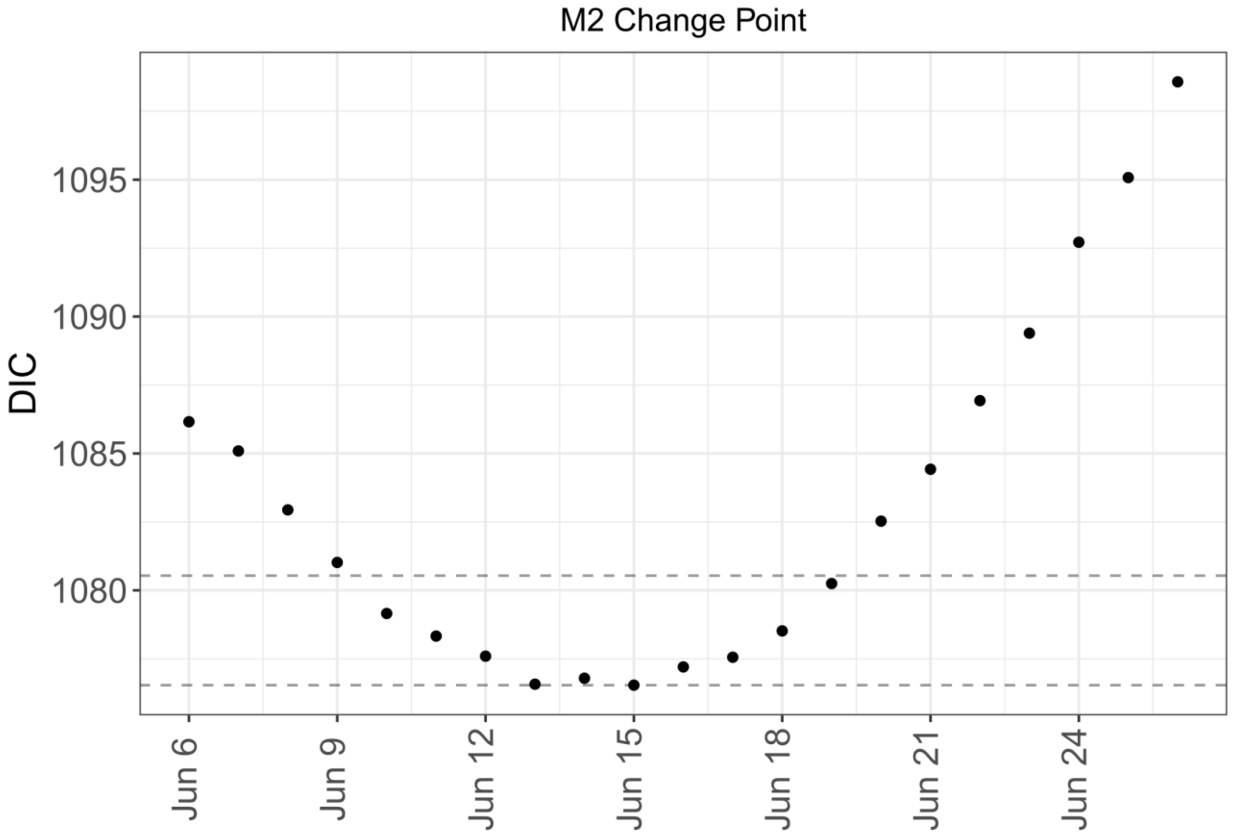

**Figure S4: Comparison between the final version of M2 and a version of M2 that includes age structure.** Solid blue lines indicate model M2 posterior means while color areas indicate 95% credible intervals. The black lines denote trajectories obtained with the model that includes age structure: these were simulated by using the parameters' posterior means obtained by calibrating M2 to data available on August 25th.

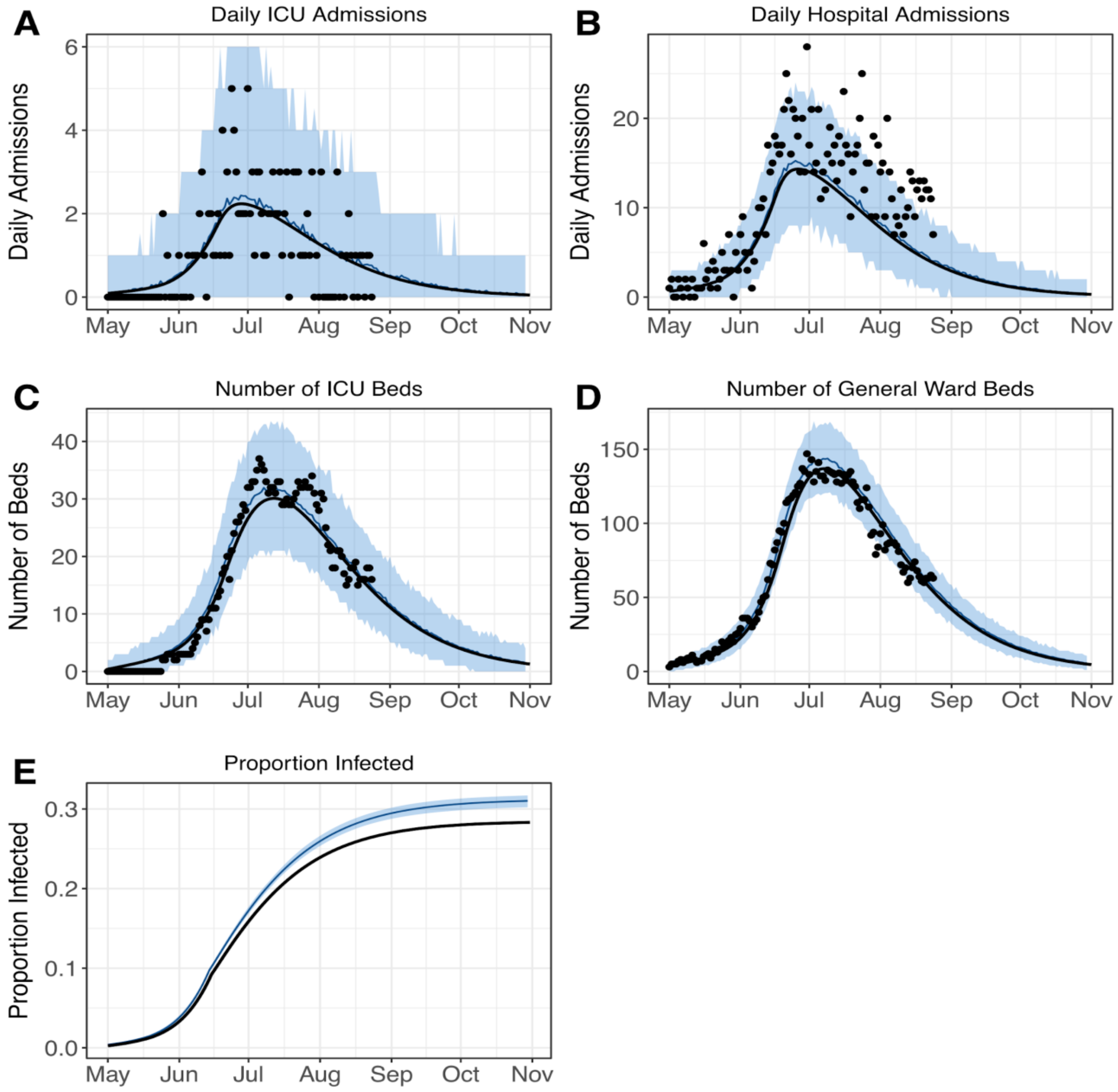

**Table S1: Key Model Parameters**

| Parameter | Model M1 | Model M2 | Model M2 (Final) |
|---|---|---|---|
| *Time spent in hospital ($\tau_H$)* | 12.8 days[b] | 11.8 days[b] | 11.8 days[b] |
| *Delay hospital to ICU admission ($\tau_{H-ICU}$)* | 1.5 days[a] | 2.6 days[b] | 2.6 days[b] |
| *Time spent in ICU ($\tau_{ICU}$)* | 11.4 days[b] | 17.6 days[b] | 15.0 [13.1, 17.4] days |
| *Probability of ICU given hospitalization ($p_{ICU}$)* | 21.5%[b] | 11.0%[b] | 15.7% [13.9%, 17.6%] |
| *Reproduction number* | 1.35 [1.26, 1.45] (before 5/20)<br><br>1.78 [1.68, 1.88] (after 5/20) | 1.40 [1.32, 1.49] (before 5/20)<br><br>1.71 [1.65, 1.77] (from 5/20 to 6/15)<br><br>1.14 [0.95, 1.31] (after 6/15) | 1.44 [1.36, 1.53] (before 5/20)<br><br>1.69 [1.65, 1.73] (from 5/20 to 6/15)<br><br>1.08 [1.07, 1.10] (after 6/15) |

[a] Value from Salje et al. 2020, estimated using data from mainland France.

[b] Values estimated using a mixture distribution as described in Lefrancq et al. 2020 and using data from French Guiana available on July 2nd 2020.

All other values were estimated with our compartmental model.
